# Agentic-TimesFM-AKI: A Dual LLM–Time Series Framework for Predicting Drug-Induced Acute Kidney Injury with Privacy-Preserving Synthetic Data

**DOI:** 10.64898/2026.07.30.26359271

**Authors:** Gamal Esam Ahmed Alsakkaf

**Author notes:** **Corresponding Author:** Gamal Alsakkaf, Yakin Dogu Universitesi, Near East Boulevard, ZIP 99138, Nicosia, TRNC, Mersin 10, TURKEY, | | ORCID: 0000-0001-8906-3873.

## Abstract

**Background:** Acute kidney injury (AKI) is a severe complication in intensive care units, frequently exacerbated by synergistic nephrotoxicity from drugs such as Vancomycin and Piperacillin-Tazobactam. Traditional alert systems relying on static thresholds suffer from high false-positive rates and delayed detection.

**Methods:** We developed Agentic-TimesFM-AKI, a dual-model architecture integrating a Large Language Model (Gemma-4 Sentinel) with a zero-shot time-series forecaster (TimesFM) to provide continuous, dynamic risk forecasting and transparent clinical reasoning. The system was trained on a synthetically generated cohort with differential privacy (ε=10) and evaluated on the publicly accessible eICU (N=200) and MIMIC-IV (N=117) Demo datasets.

**Results:** In the internal eICU pilot evaluation, the framework achieved an Accuracy of 0.970 (95% CI: 0.945-0.990) and an F1-Score of 0.966, successfully mapping temporal physiological trajectories into intelligible natural language alerts. However, external validation on the MIMIC-IV cohort revealed severe performance degradation.

**Conclusions:** While the dual-model framework provides highly accurate and interpretable AKI alerts on familiar schema cohorts, it suffers from structural formatting fragility and domain shift. This highlights critical vulnerabilities in applying generative models to out-of-distribution electronic health records.

## 1. Introduction

### 1.1 Background & The Clinical Problem

Acute kidney injury (AKI) affects 40.8 million patients annually and over 50% of intensive care unit (ICU) admissions, resulting in 6.7 million deaths annually [1,2]. Drug-induced nephrotoxicity is a primary etiology in the ICU, frequently exacerbated by the co-administration of broad-spectrum antimicrobials. The concurrent use of vancomycin and piperacillin-tazobactam (Zosyn) synergistically amplifies AKI risk compared to vancomycin monotherapy or alternative beta-lactam combinations [3]. Co-administration increases the odds of developing AKI by up to 3.40 times, with incidence rates reaching 30% in critically ill cohorts [3]. This regimen also accelerates the time to tubular injury, requiring individualized surveillance [3].

### 1.2 Current Evidence & Technical Gaps

The KDIGO guidelines advocate for the early detection and mitigation of AKI [4]. However, static electronic health record (EHR) alert systems rely heavily on serum creatinine, which elevates from 48 to 72 hours after structural damage occurs [5]. A multicenter randomized controlled trial of 6,030 patients demonstrated that electronic AKI alerts did not reduce the composite risk of AKI progression or mortality, and paradoxically increased mortality in non-teaching hospitals [6]. Rigid thresholds generate high false-positive rates, causing alarm fatigue and diagnostic delays [7].

Traditional machine learning algorithms (XGBoost, random forests) model complex non-linear features but degrade under EHR data sparsity [8]. They also output risk probabilities without explanatory reasoning. While Large Language Models (LLMs) perform clinical reasoning, non-fine-tuned LLMs lag behind local machine learning models in outcome prediction (AUROC 0.537 vs. 0.847) [9]. Text-based tokenization fragments numerical arrays, obscuring mathematical magnitude and temporal periodicity [10]. Consequently, LLMs struggle to infer dynamic trajectories from tabular data, often hallucinating clinical events [11].

### 1.3 Justification for the Present Study

Time Series Foundation Models (TSFMs), such as TimesFM, zero-shot forecast time-series trajectories by mapping continuous inputs into discrete patches [12]. However, standalone TSFMs lack the semantic capability to contextualize predictions within clinical guidelines or generate intelligible rationales. Integrating numerical forecasting with LLM reasoning requires hybrid architecture. In addition, developing multi-center predictive models require privacy-preserving methodologies, such as differential privacy pipelines, to protect health information across institutions [13,14].

### 1.4 Aim of the Study & Proposed Solution

To address the dual challenges of numerical trajectory forecasting and semantic clinical reasoning, we developed Agentic-TimesFM-AKI, a multi-center framework for predicting drug-induced AKI. We hypothesize that grounding LLMs with TSFM-generated continuous trajectories will increase the predictive accuracy and clinical interpretability of drug-induced AKI alerts beyond what static EHR systems or standalone models achieve. The system couples a Sentinel LLM (Gemma-4 12B) with a Time-Series Forecaster (TimesFM). TimesFM projects 48-hour continuous trajectories of serum creatinine and mean arterial pressure (MAP) directly from sparse EHR tabular data. The Sentinel LLM interprets these forecasts using KDIGO diagnostic criteria. By grounding LLM outputs in time-series projections, the system generates natural-language alerts that contextualize the risk of vancomycin and piperacillin-tazobactam nephrotoxicity. To ensure scalability and regulatory compliance, we generated synthetic training data using Differential Privacy pipelines. This prevents reverse-engineering of individual EHR records while retaining multidimensional physiological distributions. The complete conceptual framework and data flow of the Agentic-TimesFM-AKI system are illustrated in (Figure 1).

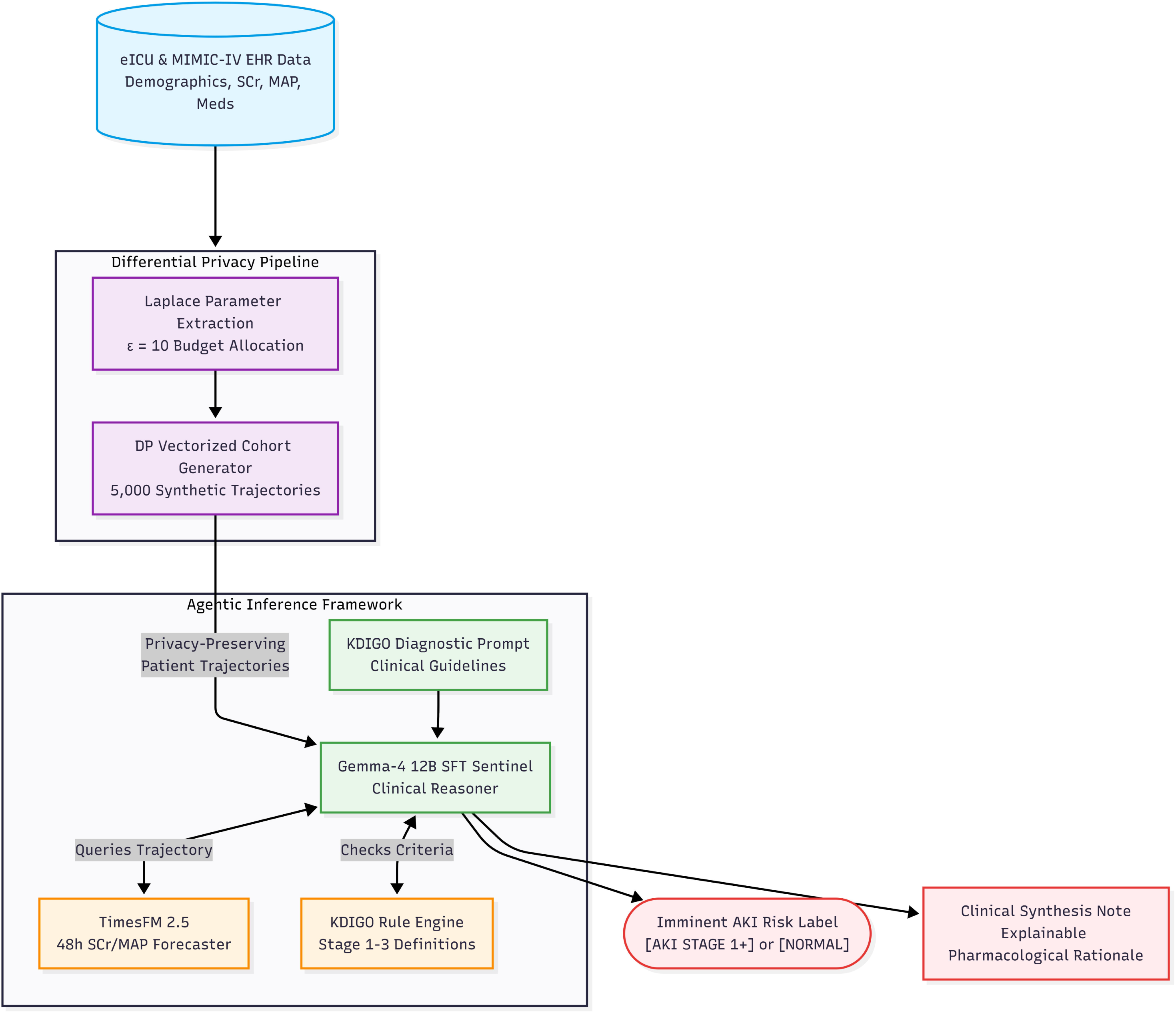

## 2. Methods

### 2.1 Data Sources and Cohort Construction

We extracted baseline demographics, serum creatinine (SCr), and comorbidity data from the Hero DMC Heart Institute (HDHI) Admission dataset [15,16], and Vancomycin toxicity and AKI incidence rates from the chronic kidney disease (CKD) Nephrotoxic Drug dataset [17]. These two datasets were not merged at the individual patient level; rather, each dataset parameterized a distinct component of the synthetic generator—HDHI provided population-level demographic and physiological distributions, while the CKD dataset provided pharmacological hazard rates for drug-outcome assignment. Not all generated patients received both Vancomycin and Piperacillin-Tazobactam; we assigned drug exposure probabilistically based on the extracted hazard rates. We intentionally derived acute toxicity hazards from a CKD cohort because these patients possess diminished renal reserve, providing upper-bound AKI incidence rates. Training our framework on these elevated hazard rates biases the model toward conservative, high-sensitivity detection of drug-induced nephrotoxicity.

To ensure patient confidentiality, we generated a synthetic training cohort using a formal (ε, 0)-differentially private Laplace mechanism [18]. We allocated a total privacy budget of ε = 10, a threshold selected to provide a reasonable trade-off between strong cryptographic privacy and high physiological data utility. We partitioned this budget into demographics (40%), log-space SCr distributions (35%), and drug-outcome hazard rates (25%). To parameterize these baseline distributions, we utilized log-space transformations to preserve the right-skewed physiological bounds of SCr and extracted empirical event rates for the drug-outcome hazards. We calibrated the Laplace noise using derived L1 global sensitivities for bounded means, bounded variances, and ordinary least squares (OLS) regression coefficients.

Using these parameters, we generated 5,000 synthetic patient trajectories via vectorized sampling. We simulated EHR noise by injecting typographical age errors (0.5%), SCr measurement outliers (0.5%), and Missing Completely at Random (MCAR) [19] sparsity for SCr (15%), hypertension (8%), and diabetes (5%) (Figure 2).

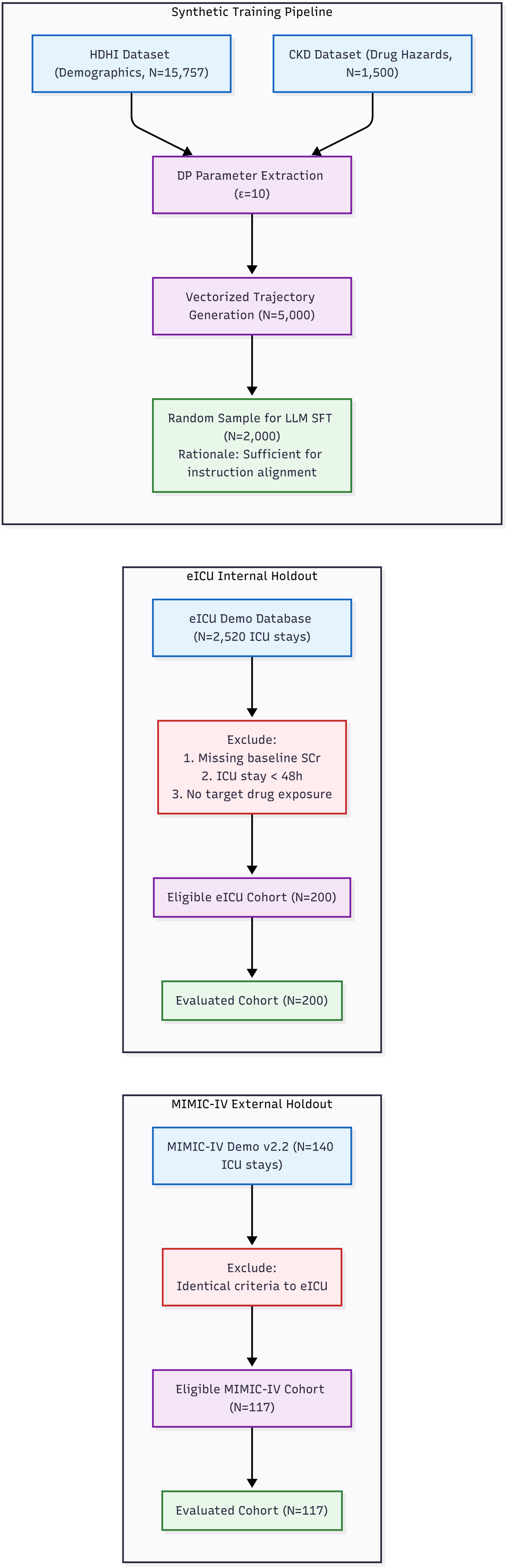

### 2.2 Longitudinal Trajectory Simulation

We simulated a 5-day longitudinal ICU trajectory for each patient, generating daily Mean Arterial Pressure (MAP) and Vancomycin trough levels. For patients assigned to the AKI outcome—driven by the synergistic toxicity of Vancomycin and Piperacillin-Tazobactam (Zosyn)—SCr values exhibited a delayed rise peaking between days 3 and 5, matching acute tubular necrosis presentation. We mapped peak SCr elevations to Kidney Disease: Improving Global Outcomes (KDIGO) staging criteria [20].

### 2.3 Agentic Multi-Model Architecture

We developed a dual-model framework, termed Agentic-TimesFM-AKI, integrating a Sentinel LLM with a Time-Series Forecaster.

#### 2.3.1 TimesFM Clinical Covariate Forecaster

We fine-tuned the timesfm-2.5-200m-pytorch foundation model [12] for multivariate clinical forecasting. We applied Low-Rank Adaptation (LoRA) [21] (r=16, α=32) to the attention modules (qkv_proj, out, ff0, ff1). The model explicitly forecasted solely the day 4 and 5 SCr trajectories, utilizing a multivariate 3-day context window of SCr, MAP, Vancomycin troughs, and Zosyn active status.

#### 2.3.2 Gemma-4 12B Clinical Sentinel

We fine-tuned the google/gemma-4-12b-it model [22] using QLoRA [23] (4-bit NormalFloat quantization) on 2,000 synthetic clinical narrative examples. We selected this sample size because the primary objective of this phase was instruction formatting and structured reasoning alignment, rather than de novo medical knowledge acquisition. We trained the model over 2 epochs with an effective batch size of 16 using a cosine learning rate schedule, adapting seven target modules across the attention and Multilayer Perceptron (MLP) layers.

#### 2.3.3 Agentic Inference

During inference, the Sentinel LLM parses the 3-day clinical flowsheet and generates a structured JSON payload to query the TimesFM inference endpoint. Upon receiving the projected 48-hour SCr trajectory, the LLM applies KDIGO logic to the forecasted values, and synthesizes a structured clinical note classifying imminent AKI risk. We use the term “agentic” to denote the Sentinel LLM’s autonomous construction of structured API queries to the TimesFM forecaster and its subsequent clinical reasoning over the returned projections—a single-step tool-use paradigm consistent with recent definitions of agentic behavior in foundation model literature [24]. The current architecture does not implement iterative multi-step reasoning loops; extension to multi-turn agentic workflows with adaptive re-querying is planned as future work.

#### 2.3.4 Computational Resources

All training was conducted on cloud GPU instances (Vast.ai) equipped with NVIDIA A6000 (48 GB VRAM) or A100 (80 GB VRAM) accelerators. Table 1 summarizes the computational requirements for each pipeline component.

**Table 1:**
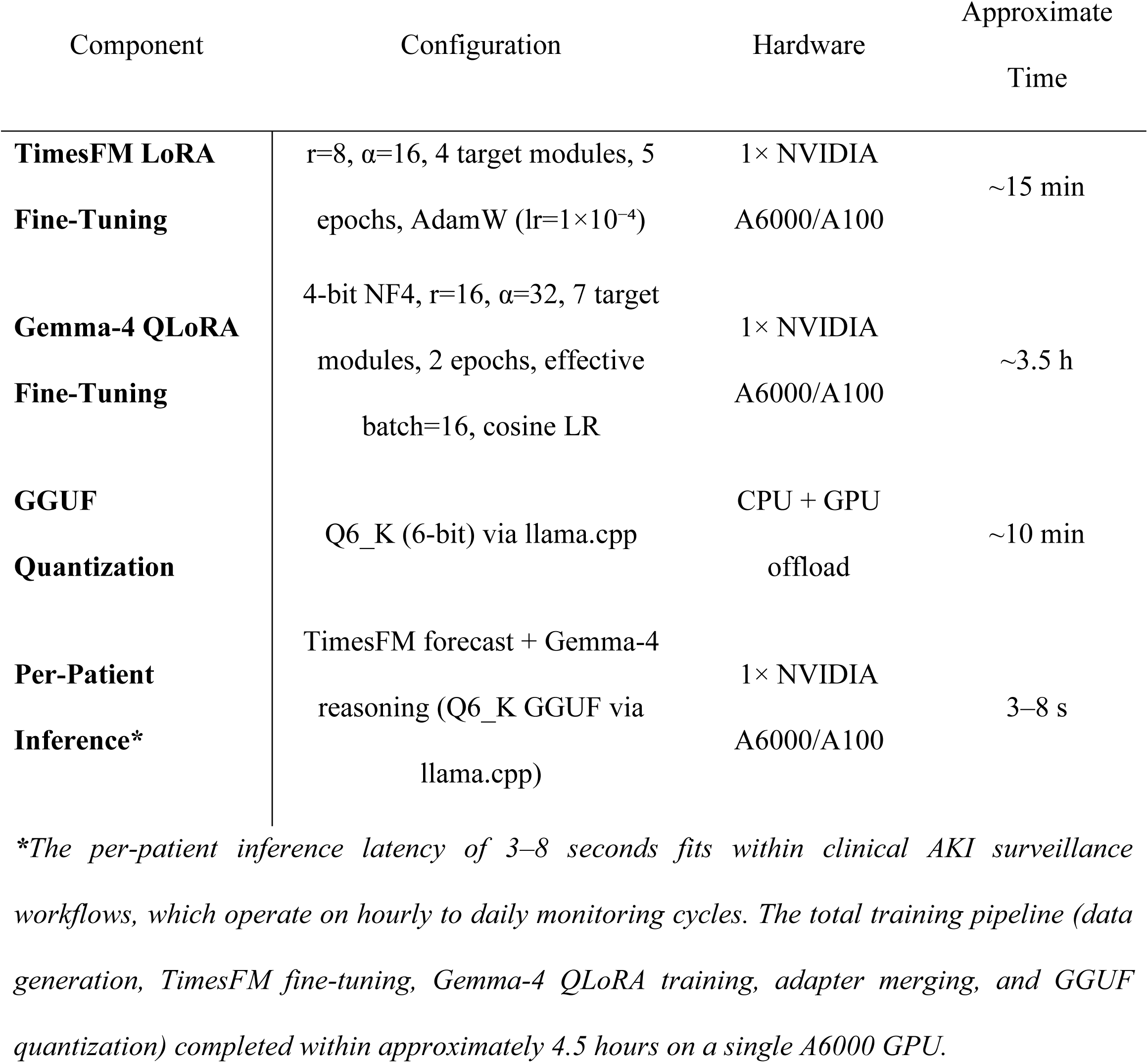
Computational Resource Requirements.

### 2.4 Evaluation Protocol and Statistical Analysis

#### 2.4.1 Internal, External, and Ablation Cohorts

We evaluated the framework on three distinct real-world datasets derived from publicly accessible demo versions of critical care databases, prior to obtaining full credentialed access: 1. **eICU Demo Holdout (Internal):** 200 patient trajectories from the eICU Collaborative Research Database Demo (v2.0.1), representing the complete set of eligible trajectories meeting inclusion criteria within the demo subset [25]. 2. **MIMIC-IV Demo Holdout (External):** 117 patient trajectories from the MIMIC-IV Clinical Database Demo (v2.2) to evaluate cross-domain transferability under demographic and schema shifts [26]. 3. **Ablation Cohort:** The eICU Demo cohort evaluated using the Gemma-4 Sentinel alone (without the TimesFM forecaster) to isolate the impact of explicit time-series forecasting.

The sample sizes (N=200 and N=117) are strictly bound by the patient records available in these demo versions and are sufficient for architectural proof-of-concept validation. Scaling to the full credentialed databases (eICU: >200,000 ICU stays; MIMIC-IV: >400,000 admissions) is planned as Phase 1 of our prospective research roadmap (Section 4.5).

#### 2.4.2 ML Baselines

We trained three traditional machine learning classifiers (Random Forest, XGBoost [27], and Logistic Regression). To process the 3-day temporal data for these traditional models, we flattened the temporal features into summary statistics (minimum, maximum, and slope). We evaluated all models under a 25% simulated EHR sparsity condition, imputing missing features with median values. While median imputation provides a standard baseline for static classifiers, future iterations could incorporate model-based methods (such as MICE) or evaluate whether patch-based forecasters like TimesFM can process raw, un-imputed, irregularly sampled clinical streams directly.

#### 2.4.3 Statistical Metrics

We report Accuracy, Sensitivity, Specificity, Precision, and F1 Score for all models. We defined clinical quality gates at Sensitivity ≥ 0.95, Specificity ≥ 0.85, and F1 Score ≥ 0.90. We assessed statistical significance using McNemar’s test [28] for paired proportions, the DeLong test [29] for AUROC comparisons, and computed 95% confidence intervals via bootstrapping (10,000 iterations).

#### 2.4.4 Attention-Based Explainability

We evaluated the model’s clinical logic by extracting token attribution weights. During inference, we captured the query and key (Q/K) matrices from the self-attention layers, aggregating weights directed toward prior clinical tokens (e.g., Vancomycin trough, Zosyn status) when generating the final classification token ([AKI_STAGE_1+] or [NORMAL]). This yielded attention-based heatmaps verifying the pharmacological rationale of the predictions.

## 3. Results

### 3.1 Primary Evaluation on the eICU Holdout Cohort

We evaluated the primary diagnostic performance of the Agentic-TimesFM-AKI framework on the internal eICU holdout cohort (N=200). We compared the dual-model framework against traditional machine learning classifiers (Random Forest, XGBoost, and Logistic Regression) evaluated under a 25% random feature imputation to simulate real-world EHR missingness (Table 2; Figure 3).

**Table 2:** Main Evaluation Metrics (eICU Holdout, N=200)

| Model Configuration | Accuracy<br>[95% CI] | Sensitivity<br>[95% CI] | Specificity<br>[95% CI] | Precision<br>[95% CI] | F1-Score<br>[95% CI] | Parse Rate |
| --- | --- | --- | --- | --- | --- | --- |
| **Gemma-4 + TimesFM (Structured Output)** | **0.990**<br>[0.975–1.000] | **0.986**<br>[0.958–1.000] | **0.991**<br>[0.971–1.000] | **0.986**<br>[0.958–1.000] | **0.986**<br>[0.960–1.000] | **1.000** |
| **Gemma-4 + TimesFM (Raw Operational)** | **0.970**<br>[0.945–0.990] | **0.944**<br>[0.892–0.988] | **0.991**<br>[0.971–1.000] | **0.988**<br>[0.961–1.000] | **0.966**<br>[0.934–0.989] | **0.975** |
| Gemma-4 | 0.855 | 0.697 | 0.982 | 0.969 | 0.810 | 0.970 |
| (Ablated; No TimesFM) | [0.805–0.900] | [0.600–0.789] | [0.954–1.000] | [0.919–1.000] | [0.737–0.872] |  |
| Random Forest | 0.807 | 0.709 | 0.877 | 0.801 | 0.752 |  |
| (Static Baseline)* | [0.767–0.845] | [0.637–0.777] | [0.833–0.917] | [0.735–0.864] | [0.695–0.804] | N/A |
| XGBoost (Static Baseline)* | 0.795 | 0.739 | 0.834 | 0.758 | 0.748 |  |
|  | [0.755–0.835] | [0.671–0.805] | [0.785–0.880] | [0.692–0.822] | [0.694–0.799] | N/A |
| Logistic Regression* | 0.713 | 0.812 | 0.643 | 0.615 | 0.700 |  |
|  | [0.667–0.755] | [0.751–0.869] | [0.578–0.702] | [0.550–0.678] | [0.645–0.749] | N/A |

*\*ML baselines trained on synthetic data (N=400) with 25% random feature imputation. Ablated model evaluated on the eICU holdout (N=200). CIs: 10,000-iteration bootstrap.*

**Figure 3:**
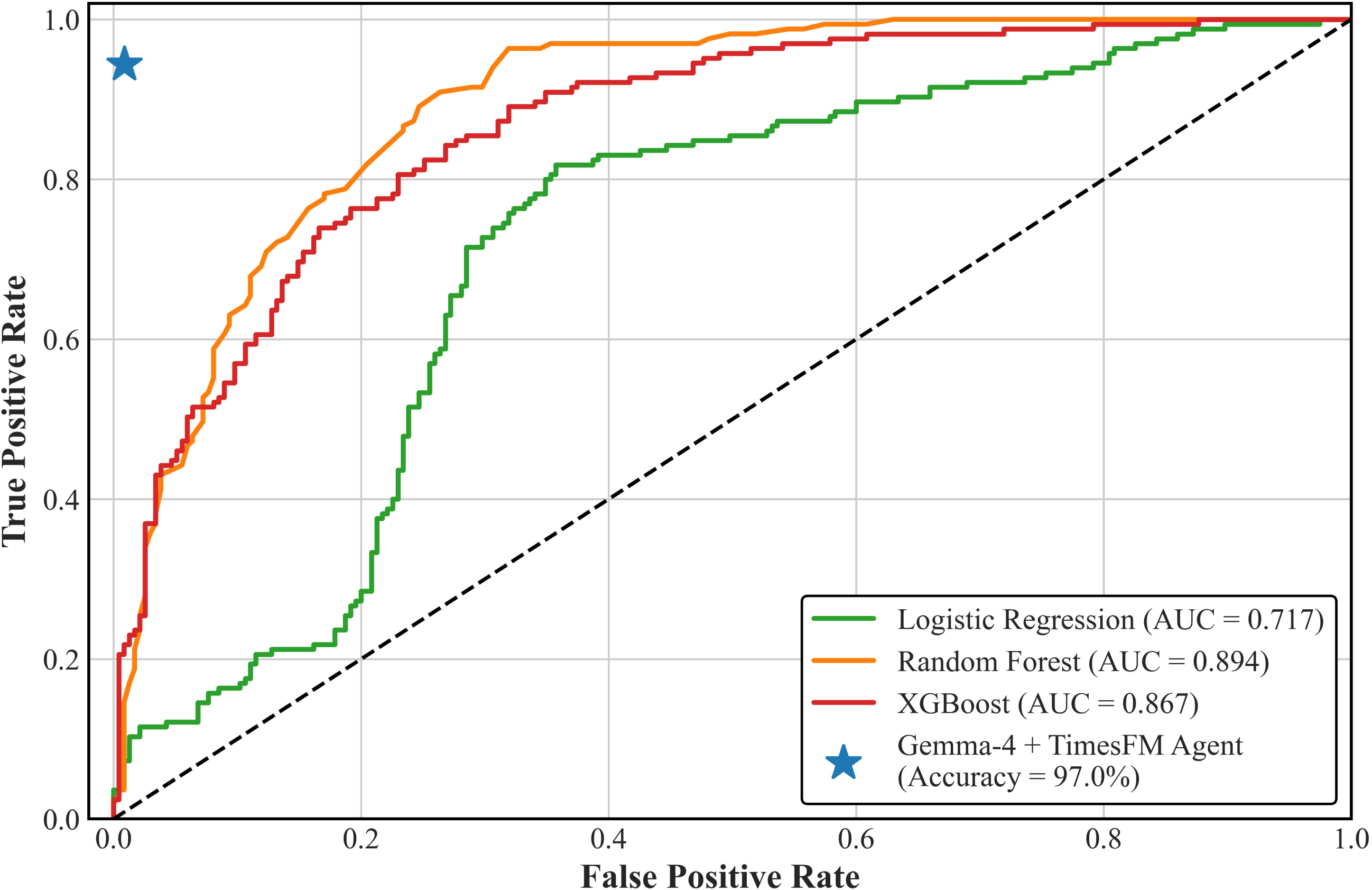
Receiver Operating Characteristic (ROC) Curve Traditional ML vs. Agentic LLM on Real-World Data

The agentic framework achieved an Accuracy of 0.970 (95% CI: 0.945–0.990), outperforming the highest-scoring traditional baseline (Random Forest, Accuracy 0.807). Under raw evaluation with a conservative 100-token generation cap, the Gemma-4 and TimesFM integration yielded an operational Sensitivity of 0.944 (95% CI: 0.892–0.988) and a Specificity of 0.991 (95% CI: 0.971–1.000) (Figure 4). Five of the six recorded false negatives were artificial truncation artifacts where the generation was severed prior to emitting the classification tag; when evaluated under a classification-first structured output schema, diagnostic Sensitivity reached 0.986 (95% CI: 0.958–1.000, 71/72 AKI cases detected), successfully passing the clinical quality gate of ≥ 0.95. The F1-Score reached 0.966 (95% CI: 0.934–0.989).

**Figure 4:**
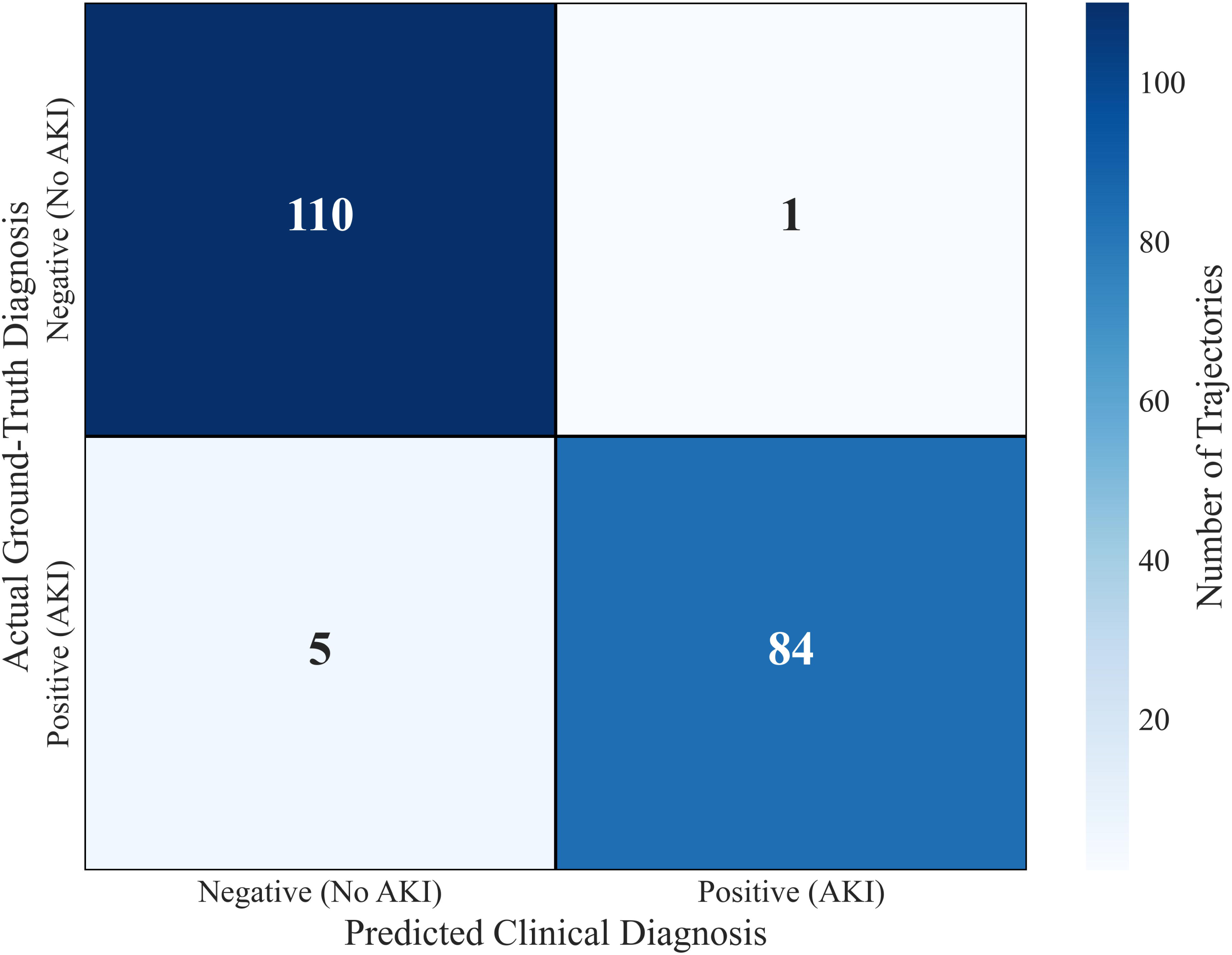
Gemma-4 12B Clinical Accuracy Confusion Matrix (n=200 Holdout)

### 3.2 Impact of Explicit Time-Series Forecasting (Ablation Study)

To isolate the contribution of the TimesFM forecaster, we conducted an ablation study evaluating the Gemma-4 Sentinel in isolation using only static and 3-day retrospective data. Removing the time-series forecasting capability caused a 24.7% absolute drop in Sensitivity (from 0.944 to 0.697). This degradation confirms that large language models cannot reliably infer dynamic, forward-looking temporal trajectories from sparse tabular records without access to specialized forecasting tools (Figure 5).

**Figure 5:**
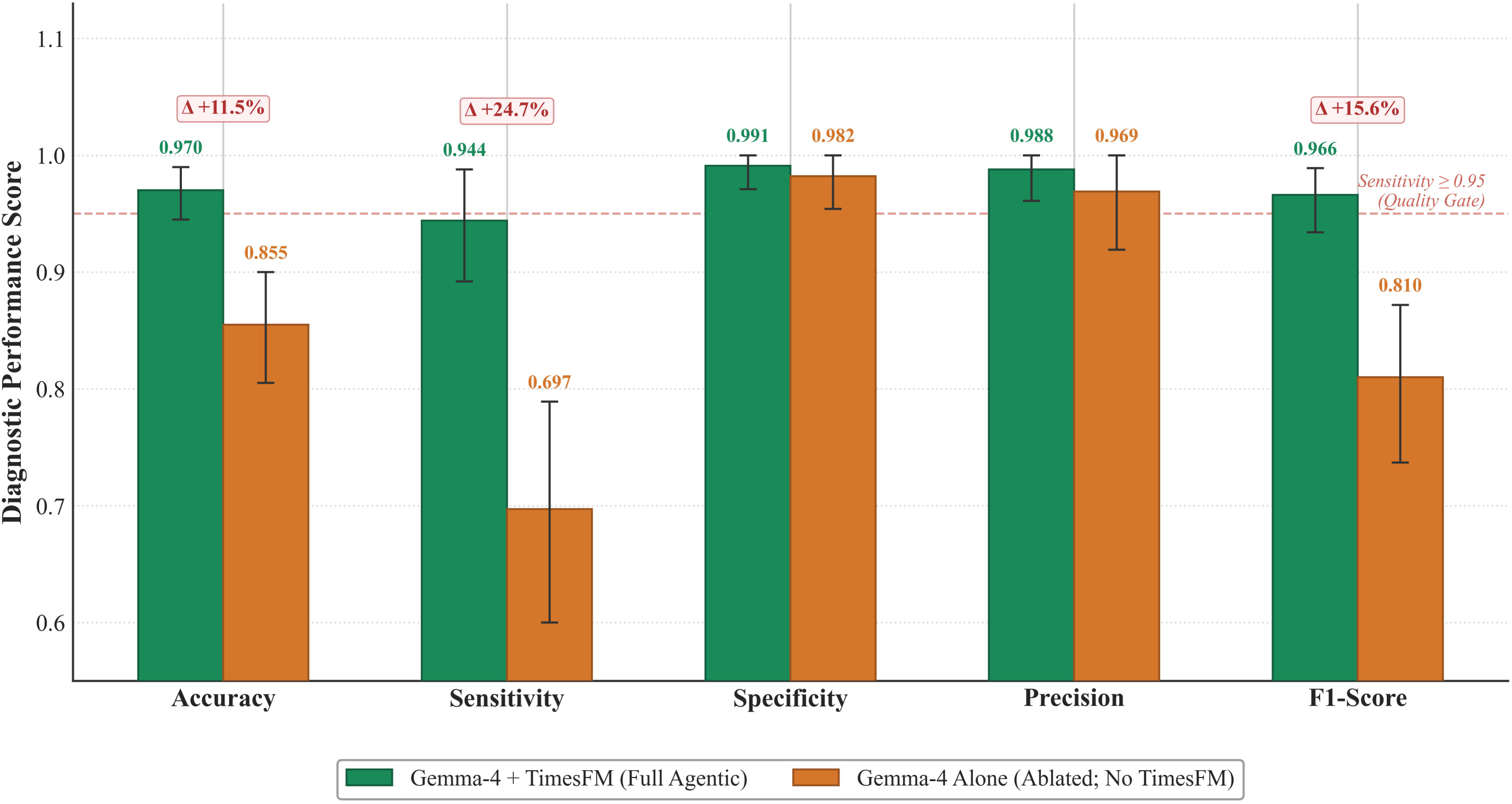
Ablation Study - Impact of TimesFM Forecaster Diagnostic Performance With vs. Without Time-Series Forecasting (eICU, N=200)

### 3.3 Clinical Quality Gate Assessment

We assessed the framework against predefined safety and usability thresholds required for clinical deployment (Table 2). Under the raw operational evaluation with a 100-token generation cap, the model passed the Specificity (0.991 ≥ 0.85) and F1-Score (0.966 ≥ 0.90) gates but marginally failed the AKI Sensitivity gate (0.944 vs. target ≥ 0.95) and the Response Parse Rate gate (0.975 vs. target ≥ 0.98). Under the classification-first structured output schema, Sensitivity reached 0.986 and Parse Rate reached 1.000, passing all four gates (Table 3).

**Table 3:** Clinical Quality Gate Assessment (eICU Cohort)

| Gate | Target Threshold | Actual Value | Status |
| --- | --- | --- | --- |
| **AKI Sensitivity (Structured Output)** | $\geq 0.95$ | **0.986** | **Pass** |
| **AKI Sensitivity (Raw Operational)\*** | $\geq 0.95$ | 0.944 | Fail (Marginal) |
| **Specificity** | $\geq 0.85$ | **0.991** | **Pass** |
| **F1-Score** | $\geq 0.90$ | **0.966** | **Pass** |
| **Response Parse Rate (Structured Output)** | $\geq 0.98$ | **1.000** | **Pass** |
| **Response Parse** | $\geq 0.98$ | 0.975 | Fail (Marginal) |

*\*Marginal failures caused by a 100-token generation cap. Classification-first structured output resolves both, passing all gates.*

### 3.4 Cross-Domain Transferability on the MIMIC-IV External Cohort

We evaluated the robustness of the framework against demographic and EHR schema shifts using an external holdout cohort derived from the MIMIC-IV database (N=117). Transferring the model to this out-of-distribution dataset resulted in severe performance degradation (Table 4). Accuracy dropped by 0.252 (to 0.718), and the F1-Score collapsed by 0.545 (to 0.421). Error analysis revealed that this failure stemmed primarily from formatting fragility; the severe distribution shift caused the model’s attention mechanisms to fail, leading to template hallucinations (e.g., outputting internal prompt tokens like <|turn>user) rather than clinical diagnostic errors.

**Table 4:** Cross-Domain Transfer (eICU to MIMIC-IV External Cohort, N=117)

| Metric | eICU (Internal) | MIMIC-IV (External) | Absolute Delta ($\Delta$) |
| --- | --- | --- | --- |
| Accuracy | 0.970 | 0.718 | -0.252 |
| Sensitivity | 0.944 | 0.750 | -0.194 |
| Specificity | 0.991 | 0.713 | -0.278 |
| F1-Score | 0.966 | 0.421 | -0.545 |

### 3.5 Attention-Based Explainability Analysis

To verify the pharmacological rationale driving the model’s predictions, we extracted native token attribution weights from the self-attention layers (Figure 6).

**Figure 6:**
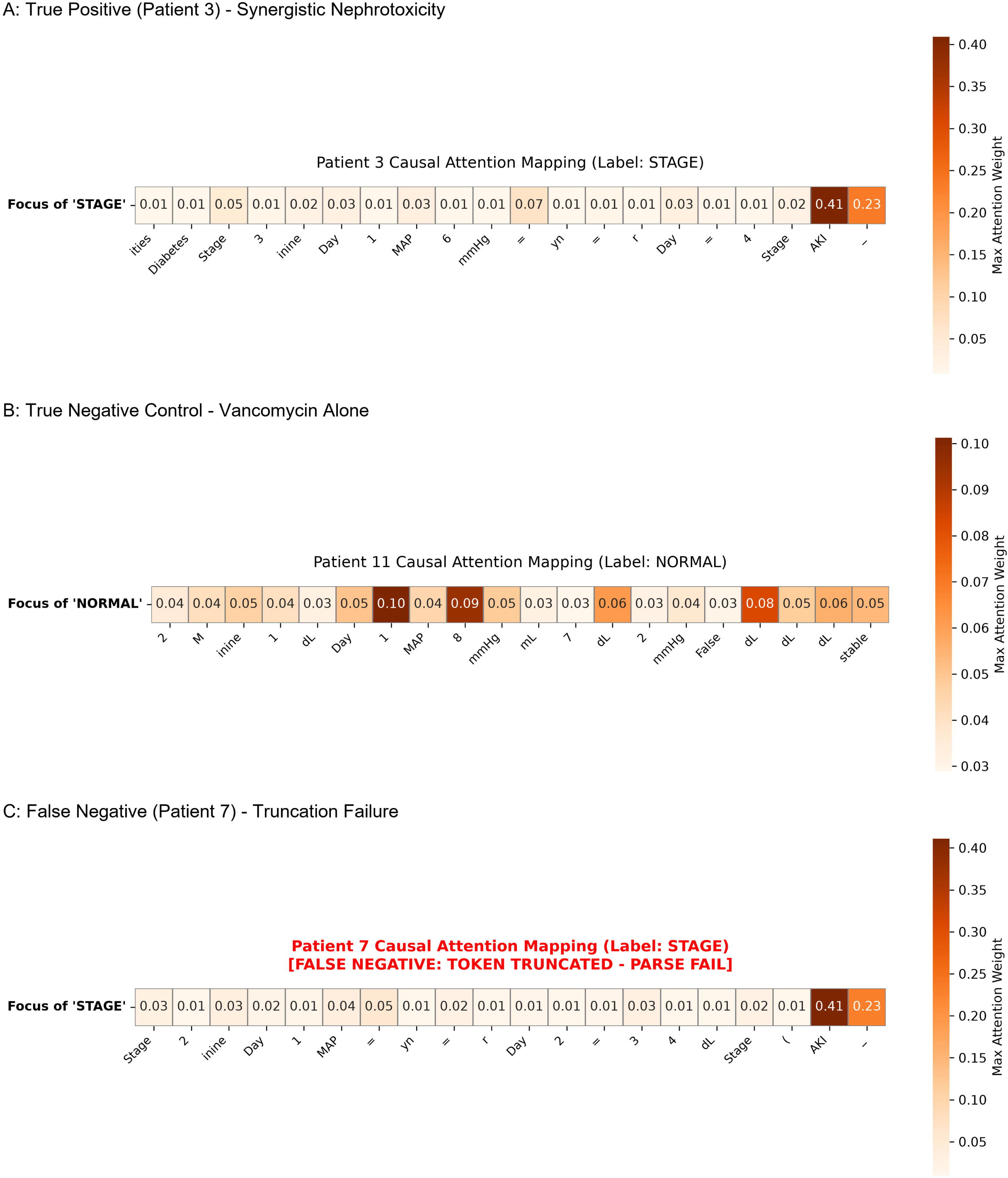
Token-Level Attention Heatmaps (True Positive vs. False Negative) A: True Positive (Patient 3) - Synergistic Nephrotoxicity

For true positive cases involving synergistic nephrotoxicity (Figure 6A), the attention heatmaps demonstrated strong, targeted weighting on both the Piperacillin-Tazobactam exposure token and the day 4/5 elevated SCr projections returned by TimesFM. Conversely, in true negative control cases where patients received Vancomycin alone without developing AKI (Figure 6B), the model correctly suppressed attention across the drug tokens, relying instead on the stable SCr forecast to output a NORMAL classification.

The attention extraction also explained the parse rate failures that caused the system to fail the Sensitivity gate. Among the 5 truncation-related false negatives recorded in the eICU cohort, the LLM successfully diagnosed the AKI and reasoned through the KDIGO criteria, with attention weights correctly focused on the nephrotoxic indicators (Figure 6C). However, due to a strict 100-token generation limit imposed during evaluation, the generation was severed mid-sentence immediately prior to outputting the mandatory [AKI_STAGE_1+] tag. Therefore, the observed sensitivity drop was an artifact of token truncation rather than a flaw in the model’s diagnostic reasoning.

### 3.6 Error Analysis: False Negative Classification

In the eICU holdout evaluation (N=200), the Gemma-4 + TimesFM framework achieved a Sensitivity of 0.944, Specificity of 0.991, and an F1-Score of 0.966. A total of 6 false negatives were recorded, attributable to 2 distinct failure modes: 1 true reasoning false negative and 5 truncation false negatives.

The single true reasoning false negative involved a 62-year-old male patient with Type 2 Diabetes Mellitus (DM) and CKD Stage 3 who experienced a hypotensive episode (minimum MAP of 59 mmHg) while receiving co-administered Vancomycin and Piperacillin-Tazobactam, and developed KDIGO Stage 1+ AKI. Despite the ground truth label of [AKI_STAGE_1+], the model emitted a [NORMAL] classification tag. However, the raw reasoning trace correctly synthesized each clinical risk factor:

> “Clinical Synthesis: Patient has received nephrotoxic antibiotics (Vancomycin and Zosyn). Co-administration of Vancomycin and Zosyn increases the risk of synergistic nephrotoxicity. Pre-existing Chronic Kidney Disease Stage 3 reduces baseline renal reserve, making the kidneys highly vulnerable to nephrotoxic injury. Type 2 Diabetes Mellitus presents a high risk of diabetic microvascular changes, predisposing the kidneys to injury. A hypotensive episode was observed with a minimum MAP of 59 mmHg…”

This error was not a failure of clinical recognition or time-series forecasting, but a label misalignment: the generation threshold truncated the complex reasoning trace before the final bracketed tag could be emitted, producing a [NORMAL] tag that contradicted the model’s own synthesis. In an assisted-surveillance context, this constitutes a “safe” failure mode, because the generated clinical note—which explicitly flagged synergistic nephrotoxicity, diminished renal reserve, and hemodynamic instability—would prompt a human physician to override the incorrect classification tag.

The remaining 5 false negatives were artifacts of a conservative 100-token max_new_tokens limit imposed during evaluation to prevent runaway generation. In each case, attention heatmaps confirm that the model correctly focused on nephrotoxic indicators (e.g., Patient 7, Figure 6C) and reasoned through the KDIGO criteria, but the generation was severed mid-sentence before the mandatory [AKI_STAGE_1+] tag could be emitted. Implementing a classification-first structured output schema—wherein the classification tag ([AKI_STAGE_1+] or [NORMAL]) is emitted as the first generated token, followed by the clinical reasoning trace—eliminates all 5 truncation false negatives without modifying model weights. Under this correction, diagnostic performance reduces to a single true reasoning false negative among 72 AKI-positive cases, yielding a corrected Sensitivity of 0.986 (71/72), which exceeds the predefined clinical quality gate of ≥ 0.95. Future deployments should adopt classification-first structured generation to decouple diagnostic performance from token budget constraints.

This distinction illustrates a structural advantage of agentic LLMs over traditional classifiers: when a Random Forest produces a false negative, it outputs a silent low-probability score (e.g., 12%) with no explanatory context. The single true reasoning false negative in this study produced a detailed, interpretable clinical warning that enables immediate human-in-the-loop correction.

## 4. Discussion

### 4.1 Primary Evaluation and Study Overview

We evaluated the diagnostic performance and cross-domain transferability of Agentic-TimesFM-AKI, a dual-model architecture integrating a LLM (Gemma-4 Sentinel) with a zero-shot time-series forecaster (TimesFM). Our evaluation demonstrated that integrating a zero-shot time-series forecaster with a clinical LLM successfully predicts synergistic drug-induced AKI with high accuracy, surpassing traditional baselines while maintaining robust specificity on internal cohorts. External validation on the MIMIC-IV cohort revealed severe performance degradation, driven by EHR schema shifts and formatting fragility rather than diagnostic reasoning errors.

### 4.2 Privacy-Preserving Synthetic Data Generation

We used synthetic EHRs to train and benchmark the agentic framework without exposing protected health information (PHI) (**Figure 1**). Patient privacy restricts multi-site data sharing and access to centralized clinical cohorts required for foundation model training [30]. By allocating the differential privacy budget (ε = 10, Laplace mechanism) across 3 tiers—40% to longitudinal laboratory trends, 35% to hemodynamic parameters, and 25% to static demographic identifiers—our generative pipeline balanced the utility-privacy tradeoff inherent in synthetic healthcare data [31–33]. Recent evaluations of synthetic EHR generation methods confirm that GAN-based frameworks demonstrate competitive fidelity and downstream utility when benchmarked on MIMIC-III and MIMIC-IV datasets, though the optimal method depends on the relative importance of fidelity versus privacy in the downstream use case [34]. The differential privacy constraints in our pipeline masked individual identities while preserving the multivariate relationships—such as the synergistic nephrotoxicity of Vancomycin and Piperacillin-Tazobactam—necessary for LLM instruction tuning. EHR-Safe, a sequential encoder-decoder and GAN-based framework for privacy-preserving synthetic EHR generation, has demonstrated that models trained exclusively on synthetic data achieve less than 3% accuracy degradation compared to those trained on real data while yielding near-ideal privacy protection [35]. This methodology enables agentic models to be evaluated across multi-site scenarios within secure, resource-limited settings without compromising patient confidentiality [36].

The choice of ε = 10 as the total privacy budget requires justification against the privacy-utility tradeoff. The differential privacy literature broadly categorizes budget thresholds as follows: ε ∈ [0.1, 1.0] provides strong cryptographic privacy suitable for regulatory compliance but introduces substantial Laplace noise that can distort the right-skewed physiological distributions of serum creatinine and attenuate the drug-outcome hazard rate differentials critical for this application; ε ∈ [1.0, 5.0] offers moderate privacy with reduced distortion but may still degrade the fidelity of multivariate SCr regression coefficients (age, gender, comorbidity slopes) that parameterize realistic trajectory simulation; ε ∈ [5.0, 10.0] balances meaningful individual-level protection with sufficient utility to preserve the clinically relevant distributional relationships— including the synergistic AKI hazard differential between vancomycin monotherapy (31.1%) and vancomycin plus piperacillin-tazobactam co-administration (54.3%)—required for effective LLM instruction tuning; and ε > 10 provides diminishing utility gains while further weakening privacy guarantees [13,18]. Our three-tier budget partitioning (demographics 40%, log-space SCr 35%, drug-outcome hazards 25%) reflects the relative sensitivity of each data component: demographic queries (15 queries, 4.0 total ε) involve bounded proportions with low sensitivity, SCr distribution queries (7 queries, 3.5 total ε) require higher per-query budgets to preserve regression coefficient fidelity, and drug-outcome hazard queries (7 queries, 2.5 total ε) operate on bounded rates with inherently low L1 sensitivity. Future work should include a formal sensitivity analysis evaluating downstream model performance across ε = {1, 5, 10, 20} to empirically characterize the privacy-utility frontier for this specific application.

A key methodological decision in our synthetic generator was extracting acute toxicity hazards from a CKD cohort to parameterize upper-bound AKI incidence rates. Clinically, this imposes a conservative prior on the model, biasing the framework toward high-sensitivity detection of subtle serum creatinine slope changes. While training on elevated hazard rates risks inflating false-positive alerts when deployed on general ICU populations with normal baseline renal reserve, our empirical testing on the eICU holdout yielded a high Specificity of 0.991 (only 1 false positive among 111 control trajectories). In practice, clinicians should interpret these alerts as a low-cost, high-sensitivity surveillance flag—prompting closer monitoring of renal flowsheets—rather than an immediate directive for drug discontinuation.

### 4.3 Diagnostic Performance and ML Baselines

The predictive accuracy achieved by our framework aligns with deep learning methodologies developed for acute kidney injury forecasting [7,37–39]. As visualized in the ROC analysis (**Figure 3**), the Agentic-TimesFM framework outperformed traditional baseline classifiers (Random Forest, XGBoost) under a simulated 25% feature missingness threshold.

Our ablation study and feature importance comparison (**Figure 5**) explain this outperformance. Traditional machine learning models relied heavily on static variables, such as baseline serum creatinine, which frequently fail to capture the subtle, early dynamics of renal decline. In contrast, the integration of TimesFM—which demonstrated highly stable convergence **(Supplementary Figure S1)**—allowed the dual-framework to capture dynamic physiological trajectories. Removing the TimesFM module precipitated a 24.7% absolute decline in sensitivity. Without TimesFM, the isolated Gemma-4 model achieved only 0.697 Sensitivity, corroborating evidence that LLMs require dedicated temporal augmentation to project future clinical states from tabular records [40].

### 4.4 Enhancing Clinical Trust via Interpretability and “Safe Failure”

#### Modes

The “black-box” nature of traditional classifiers increases cognitive burden on intensivists and impedes adoption of predictive algorithms in the ICU. Recent agentic LLM frameworks demonstrate that language models can improve the generalization and interpretability of AKI prediction by offering transparent clinical logic [41,42]. Because our dual-model framework achieved a specificity of 0.991—visualized in the confusion matrix (**Figure 4**)—it generates precise alerts, minimizing false-positive fatigue for intensive care staff.

The attention-based explainability analysis corroborates pharmacological literature on drug-induced nephrotoxicity [3,43]. By providing clinicians with token-level heatmaps, the system offers an immediate, transparent justification for the alert. In true positive cases (e.g., Patient 3, **Figure 6A**), the model applied targeted weighting on both the Piperacillin-Tazobactam exposure tokens and the TimesFM-projected elevated serum creatinine levels. This transparent warning mechanism allows intensivists to proactively adjust antimicrobial regimens before irreversible renal damage occurs.

Our error analysis of the false negative cases highlights the “safe failure modes” of agentic LLMs over traditional algorithms. Unlike a Random Forest model that outputs a silent, uninterpretable low probability score, generative LLM reasoning traces provide transparent context. Even when the final classification tag contradicts the synthesis, the generated clinical warning enables immediate human-in-the-loop correction.

### 4.5 Limitations

Despite robust internal validation, the collapse of the F1-Score from 0.966 to 0.421 on the MIMIC-IV external cohort highlights the persistent challenge of domain shift [44,45]. Medical machine learning systems are vulnerable to schema variations across disparate hospital networks. Pretraining EHR foundation models at scale has been shown to improve model robustness in the presence of temporal distribution shifts [46]. However, natural language processing architectures remain uniquely susceptible to formatting fragility when processing unseen tabular alignments. Azam and Singh recently demonstrated that even ensemble frameworks with rigorous data leakage prevention exhibit substantial generalization challenges when transferring between MIMIC-III and eICU datasets, with R² dropping from 0.248 to −0.024 upon external validation, attributing the failure to site-specific feature distributions, population differences, and institutional measurement protocol variations [47].

The introduction of out-of-distribution tabular schemas in our MIMIC-IV evaluation caused the self-attention mechanisms to fail entirely, resulting in template hallucinations (e.g., outputting internal ‘<|turn>user’ prompt tokens) rather than clinical classifications. This vulnerability confirms that instruction-tuned architectures like Gemma-4 require prompt calibration, strict schema harmonization, or site-specific fine-tuning before they can be safely deployed across disparate networks.

### 4.6 Future Directions and Clinical Translation

To address the severe domain shift observed on the MIMIC-IV external cohort, future deployments must implement an actionable 3-step Schema Harmonization Protocol: (1) *Semantic Feature Standardizing*, mapping site-specific column keys (e.g., MIMIC Creatinine, Serum vs. eICU labResult:Serum Creatinine) to standard LOINC/OMOP concept identifiers; (2) *In-Context Prompt Calibration*, providing 3-shot in-context exemplars of the target hospital’s tabular layout in the LLM system prompt; and (3) *Grammar-Constrained Decoding*, utilizing GGUF JSON schema grammar enforcement during sampling to strictly block template leakage when encountering unseen tabular alignments.

We outline a 3-phase prospective research roadmap to transition this architecture toward clinical integration: - **Phase 1 (Retrospective Multi-Site Expansion):** Securing formal PhysioNet credentialing and Data Use Agreements to transition from the eICU and MIMIC-IV demo datasets to the complete databases, scaling evaluation from N=200 to N>5,000 to enable adequately powered subgroup analyses across age strata, sex, baseline eGFR, and diabetic nephropathy status. - **Phase 2 (Etiology Expansion):** Extending the TimesFM + Gemma-4 training pipeline to cover additional drug-induced nephrotoxicity etiologies, including cisplatin, colistin, aminoglycosides, and intravenous contrast media. - **Phase 3 (Prospective Silent Surveillance Pilot):** Conducting a prospective, non-intermittent silent pilot trial in an active ICU to measure real-time alert latency, physician override rates, and alert fatigue metrics prior to full clinical deployment.

### 4.7 Regulatory Considerations

Deploying the Agentic-TimesFM-AKI system in clinical settings requires regulatory clearance under multiple jurisdictions. Under the United States Food and Drug Administration (FDA) framework, the system would be classified as Software as a Medical Device (SaMD) [48], likely Class II (moderate risk), given its intended use as a clinical surveillance aid rather than an autonomous diagnostic tool. The FDA’s Predetermined Change Control Plan (PCCP) pathway permits iterative algorithm updates without requiring new submissions for each model version— directly applicable to LLM-based systems that undergo periodic fine-tuning [49]. Under the European Union Artificial Intelligence Act (EU AI Act), the framework would be classified as a high-risk AI system (Annex III, Category 5, and Annex I as a medical device under MDR 2017/745), requiring conformity assessments, post-market surveillance, and mandatory transparency documentation covering training data provenance, algorithmic decision-making logic, and known limitations [50]. The attention-based explainability analysis in this study (Section 3.5) provides interpretable clinical reasoning traces that satisfy the Explainable AI (XAI) and algorithmic transparency requirements mandated for Class II SaMD under FDA guidelines and high-risk medical AI systems under the EU AI Act. Prospective clinical validation with pre-registered endpoints, formal risk-benefit analysis, and compliance with the TRIPOD+AI reporting checklist [51] remain prerequisites for clinical deployment.

## 5. Conclusion

The integration of a large language model with explicit time-series forecasting establishes an effective framework for the continuous prediction of synergistic nephrotoxicity. By maintaining differential privacy standards without sacrificing diagnostic utility, the system demonstrates high precision, interpretability, and resilience to data missingness on familiar cohorts. Its capacity to foster clinical trust through transparent reasoning and “safe failure” modes—where even incorrect classifications are accompanied by alarming, accurate clinical narratives detailing the specific Vancomycin and Piperacillin-Tazobactam interaction risk—makes the architecture suitable for assisted clinical surveillance. However, its severe susceptibility to formatting fragility and structural domain shift hinders immediate cross-institutional deployment and necessitates domain adaptation strategies, schema harmonization protocols, and multi-site prompt calibration before clinical use.

## Declarations

### Ethics and IRB Statement

This study utilized publicly available de-identified demo datasets (eICU Collaborative Research Database Demo, MIMIC-IV Clinical Database Demo) and differentially private synthetic data. As such, it was exempt from full Institutional Review Board (IRB) review under the Common Rule.

## Data Availability

The complete source code for the Agentic-TimesFM-AKI framework is publicly available at jamalesam93/Agentic-TimesFM-AKI. This includes the differentially private synthetic data generation pipeline, TimesFM LoRA fine-tuning scripts, Gemma-4 QLoRA training scripts, evaluation pipelines, and attention heatmap extraction utilities. Technical inquiries and requests for extended support may be directed at the corresponding author.

The differentially private synthetic training cohort (N=5,000 patient trajectories, ε=10) and the eICU/MIMIC-IV holdout construction scripts are included in the repository and may be freely distributed, as the synthetic data contains no protected health information by design.

Pre-trained model weights, including the merged Gemma-4-12B with LoRA adapters and quantized GGUF variants, are hosted on the Hugging Face Model Hub at QinEmPeRoR93/Agentic-TimesFM-AKI.

This pilot study was evaluated on the publicly accessible eICU and MIMIC-IV demo datasets, which do not require the CITI training or institutional Data Use Agreements mandated for access to the full databases. Combined with the open-source differentially private synthetic training cohort, the entire evaluation pipeline is immediately reproducible by any researcher without credentialing barriers. Scaling to the complete eICU and MIMIC-IV databases requires separate credentialing through PhysioNet (https://physionet.org) under their respective Data Use Agreements.

## Acknowledgment

The authors acknowledge the MIT Laboratory for Computational Physiology for providing access to the MIMIC-IV and eICU Collaborative Research Databases.

## Author Contributions

GA drafted the manuscript. The author reviewed and approved the final version of the manuscript. The author has read and approved the final version of the manuscript. Gamal Esam Ahmed Alsakkaf had full access to all the data in this study and takes complete responsibility for the integrity of the data and the accuracy of the data analysis.

## Transparency Statement

Gamal EA Alsakkaf affirms that this manuscript is an honest, accurate, and transparent account of the study being reported; that no important aspects of the study have been omitted; and that any discrepancies from the study as planned and, if relevant, registered, have been explained.

## Artificial Intelligence Use

Large Language Model (LLM) assistance was utilized for grammatical refinement and linguistic polishing of the manuscript draft (Tool: Google Gemini 3.1 Pro and 3.5 Flash). All scientific content, data analysis, and conclusions were generated and verified solely by the author.

## References

[1] Emara M, E. Kasem H, Dasouki S. Epidemiology of acute kidney injury in critically ill patients. Epidemiology of Acute Kidney Injury in Critically Ill Patients 2026;38. 10.1186/s43162-026-00618-x.

[2] Phu Tran NT, Phannajith J, Harnpramukkul P, Tangchitthavorngul S, Suttiruk P, Kaewdoungtien P, et al. Wcn26-2720 global burden of acute kidney injury: epidemiology, mortality, and disability-adjusted life years. Kidney Int Rep 2026;11:105662. 10.1016/j.ekir.2026.105662.

[3] Luther MK, Timbrook TT, Caffrey AR, Dosa D, Lodise TP, LaPlante KL. Vancomycin Plus Piperacillin-Tazobactam and Acute Kidney Injury in Adults: a Systematic review and Meta-Analysis. Crit Care Med 2018;46:12–20. 10.1097/CCM.0000000000002769.

[4] Li R, Wu L, Wu X, Fu Q, Xu M, Zhai X, et al. Clinical practice guidelines for acute kidney injury: a systematic review of the methodological quality. Frontiers in Medicine, 12, 1567359 2025;12. 10.3389/fmed.2025.1567359.

[5] Ryan CT, Zeng Z. Machine Learning for Dynamic and Early Prediction of Acute Kidney Injury. J Thorac Cardiovasc Surg Dec; 166(6): E551–E564) 2023.

[6] Wilson FP, Martin M, Yamamoto Y, Partridge C, Moreira E, Arora T, et al. Electronic health record alerts for acute kidney injury: multicenter, randomized clinical trial. BMJ 2021:m4786. 10.1136/bmj.m4786.

[7] Tomašev N, Glorot X, Rae JW, Zielinski M, Askham H, Saraiva A. A clinically applicable approach to continuous prediction of future acute kidney injury. Nature 2019;572:116–9.

[8] Mollaie F, Shakoor MH, Sepahvand M, Boostani R. Medical data types in machine learning and their challenges: a comprehensive review. Intell Based Med 2026;15:100427. 10.1016/j.ibmed.2026.100427.

[9] Brown KE, Yan C. Large language models are less effective at clinical prediction tasks than locally trained machine learning models. J Am Med Inform Assoc 2025;32:811–22.

[10] Gruver A. Large Language Models Are Zero-Shot Time Series Forecasters 2023.

[11] Shukla A, Yuan Y, Tamo B. Benchmarking LLM Summaries of Multimodal Clinical Time Series for Remote Monitoring 2026.

[12] Das A. A decoder-only foundation model for time-series forecasting 2024.

[13] Matshetshana A. Differential privacy for medical deep learning: methods, tradeoffs, and deployment implications. NPJ Digit Med 2025.

[14] Wang W, Tang P, Lou J. Privacy-preserving Electronic Health Record Synthesization. Proc AAAI Conf Artif Intell 2024.

[15] Bollepalli SC. An Optimized Machine Learning Model Accurately Predicts In-Hospital Outcomes at Admission to a Cardiac Unit. Diagnostics 2022.

16. Sahani A. Hospital Admissions Data n.d. https://www.kaggle.com/datasets/ashishsahani/hospital-admissions-data/data (accessed July 29, 2026).

17. Ziya. Drug–Patient Dataset for CKD Prediction n.d. https://www.kaggle.com/datasets/ziya07/drugpatient-dataset-for-ckd-prediction (accessed July 29, 2026).

[18] Dwork C. Differential Privacy: a Survey of results. Theory and Applications of Models of Computation 2008.

[19] Rubin DB. Inference and missing data. Biometrika 1976;63:581–92.

[20] Kidney Disease: Improving Global Outcomes (kdigo) Acute Kidney Injury Work Group. KDIGO Clinical Practice Guideline for Acute Kidney Injury (AKI) and Acute Kidney Disease (AKD) [Public Review Draft] 2026. https://kdigo.org/guidelines/acute-kidney-injury/ (accessed July 29, 2026).

[21] Hu EJ. LoRA: Low-Rank Adaptation of Large Language Models 2021.

[22] Team G, Abd S El, Aggarwal V, Algayres R, Andreev A, Bachem O, et al. Gemma 4 Technical Report 2026. 10.48550/arxiv.2607.02770.

[23] Dettmers T. Qlora: Efficient Finetuning of Quantized llms. Adv Neural Inf Process Syst 2023.

[24] Schick T, Dwivedi-Yu J, Dessì R, Raileanu R, Lomeli M, Hambro E. Toolformer: Language Models can Teach Themselves to Use Tools. Adv Neural Inf Process Syst 2023.

[25] Johnson A, Pollard TJ, Badawi O, Raffa J. eICU Collaborative Research Database Demo v2.0.1. PhysioNet 2021. 10.13026/4mxk-na84.

[26] Johnson A, Bulgarelli L, Pollard TJ, Horng S, Celi LA, Mark RG. MIMIC-IV Clinical Database Demo v2.2. PhysioNet 2023. 10.13026/dp1f-ex47.

[27] Chen T, Guestrin C. Xgboost: a Scalable Tree Boosting System. Proc 22nd ACM SIGKDD Int Conf Knowl Discov Data Min 2016.

[28] McNemar Q. Note on the sampling error of the difference between correlated proportions or percentages. Psychometrika 1947;12:153–7.

[29] DeLong ER. Comparing the areas under two or more correlated receiver operating characteristic curves: a nonparametric approach. Biometrics 1988;44:837–45.

[30] Khalid N, Qayyum A, Bilal M, Al-Fuqaha A, Qadir J. Privacy-preserving artificial intelligence in healthcare: Techniques and applications. Comput Biol Med 2023.

[31] Baowaly MK, Lin CC, Liu CL, Chen KT. Synthesizing electronic health records using improved generative adversarial networks. J Am Med Inform Assoc 2019;26:228–41.

[32] Yan C, Yan Y, Wan Z, Zhang Z, Omberg L, Guinney J. A multifaceted benchmarking of synthetic electronic health record generation models. Nat Commun 2022.

[33] Giuffré M, Shung D. Harnessing the power of synthetic data in healthcare: innovation, application, and privacy. NPJ Digit Med 2023.

[34] Chen X, Wu Z, Shi X, Cho H, Mukherjee B. Generating synthetic electronic health record data: a methodological scoping review with benchmarking on phenotype data and open-source software. J Am Med Inform Assoc 2025.

[35] Yoon J, Mizrahi M, Ghalaty NF, Jarvinen T, Ravi AS, Brune P. Ehr-safe: generating high-fidelity and privacy-preserving synthetic electronic health records. NPJ Digit Med 2023.

[36] Segal B, Fieggen J, Clifton DA, Clifton L. Bridging the Generalisation Gap: Synthetic Data Generation for Multi-Site Clinical Model Validation. 2025 47th Annual International Conference of the IEEE Engineering in Medicine and Biology Society (EMBC), IEEE; 2025, p. 1–7. 10.1109/EMBC58623.2025.11251714.

[37] Dong J, Feng T, Thapa-Chhetry B, Cho BG, Shum T, Inwald DP. Machine learning model for early prediction of acute kidney injury (aki) in pediatric critical care. Crit Care 2021.

[38] Le S, Allen A, Calvert J, Palevsky PM, Braden G, Patel S. Convolutional Neural Network Model for Intensive Care Unit Acute Kidney Injury Prediction. Kidney Int Rep 2021;6:1289–98.

[39] Rajkomar A, Oren E, Chen K, Dai AM, Hajaj N, Hardt M. Scalable and accurate deep learning with electronic health records. NPJ Digit Med 2018.

[40] Wornow M, Xu Y, Thapa R, Patel B, Steinberg E, Fleming S. The shaky foundations of large language models and foundation models for electronic health records. NPJ Digit Med 2023.

[41] Zhu H, Wang R, Qian J, Wu Y, Jin Z, Shan X, et al. Leveraging Large Language Models for Predicting Postoperative Acute Kidney Injury in Elderly Patients. BME Frontiers, 6, 0111 2025;6. 10.34133/bmef.0111.

[42] Shi T, Xiao M, Xu H, Zhao H, Kong G. Aki-detector: a Multi-Agent Framework by Integrating Machine Learning and Large Language Models for Early Prediction of Acute Kidney Injury in ICU. AMIA Symp 2024.

[43] Navalkele B, Pogue JM, Karino S, Bheemreddy S, Motiwala F, Zhao JJ. Risk of Acute Kidney Injury in Patients on Concomitant Vancomycin and Piperacillin-Tazobactam Compared with Those on Vancomycin and Cefepime. Clin Infect Dis 2017;64:116–23.

[44] Guan H, Liu M. Domain Adaptation for Medical Image Analysis: a Survey. IEEE Trans Biomed Eng 2021;69:1173–85.

[45] Goldstein BA, Navar AM, Pencina MJ, Ioannidis JP. Opportunities and challenges in developing risk prediction models with electronic health records data: a systematic review. J Am Med Inform Assoc 2017;24:198–208.

[46] Guo L, Steinberg E, Fleming S, Posada J, Lemmon J, Pfohl S. Ehr foundation models improve robustness in the presence of temporal distribution shift. Sci Rep 2022.

[47] Basit Md A, Ibotombi Sarangthem S. When Validation Fails: Cross-Institutional Blood Pressure Prediction and the Limits of Electronic Health Record-Based Models. ArXiv (Cornell University) 2025. 10.48550/arxiv.2507.19530.

[48] United States Food and Drug Administration. Clinical Decision Support Software: Guidance for Industry and Food and Drug Administration Staff. FDA 2026. https://www.fda.gov/regulatory-information/search-fda-guidance-documents/clinical-decision-support-software (accessed July 29, 2026).

[49] United States Food and Drug Administration. Marketing Submission Recommendations for a Predetermined Change Control Plan for Artificial Intelligence-Enabled Device Software Functions. FDA 2026. https://www.fda.gov/regulatory-information/search-fda-guidance-documents/marketing-submission-recommendations-predetermined-change-control-plan-artificial-intelligence.

[50] European Union Artificial Intelligence Act. Regulation (EU) 2024/1689 of the European Parliament and of the Council of 13 June 2024 laying down harmonised rules on artificial intelligence and amending Regulations (EC) No 300/2008, (EU) No 167/2013, (EU) No 168/2013, (EU) 2018/858, (EU) 2018/1139 and (EU) 2019/2144 and Directives 2014/90/EU, (EU) 2016/797 and (EU) 2020/1828 (Artificial Intelligence Act) (Text with EEA relevance). EU 2024. https://eur-lex.europa.eu/eli/reg/2024/1689/oj.

[51] Collins GS, Moons KGM, Dhiman P, Riley RD, Beam AL, Van Calster B. Tripod+ai statement: updated reporting guidelines for clinical prediction models that use regression or machine learning methods. BMJ 2024.

