## Supplementary figures and images for "Agentic-TimesFM-AKI: A Dual LLM–Time Series Framework for Predicting Drug-Induced Acute Kidney Injury with Privacy-Preserving Synthetic Data"

### Supplemental Figure 1

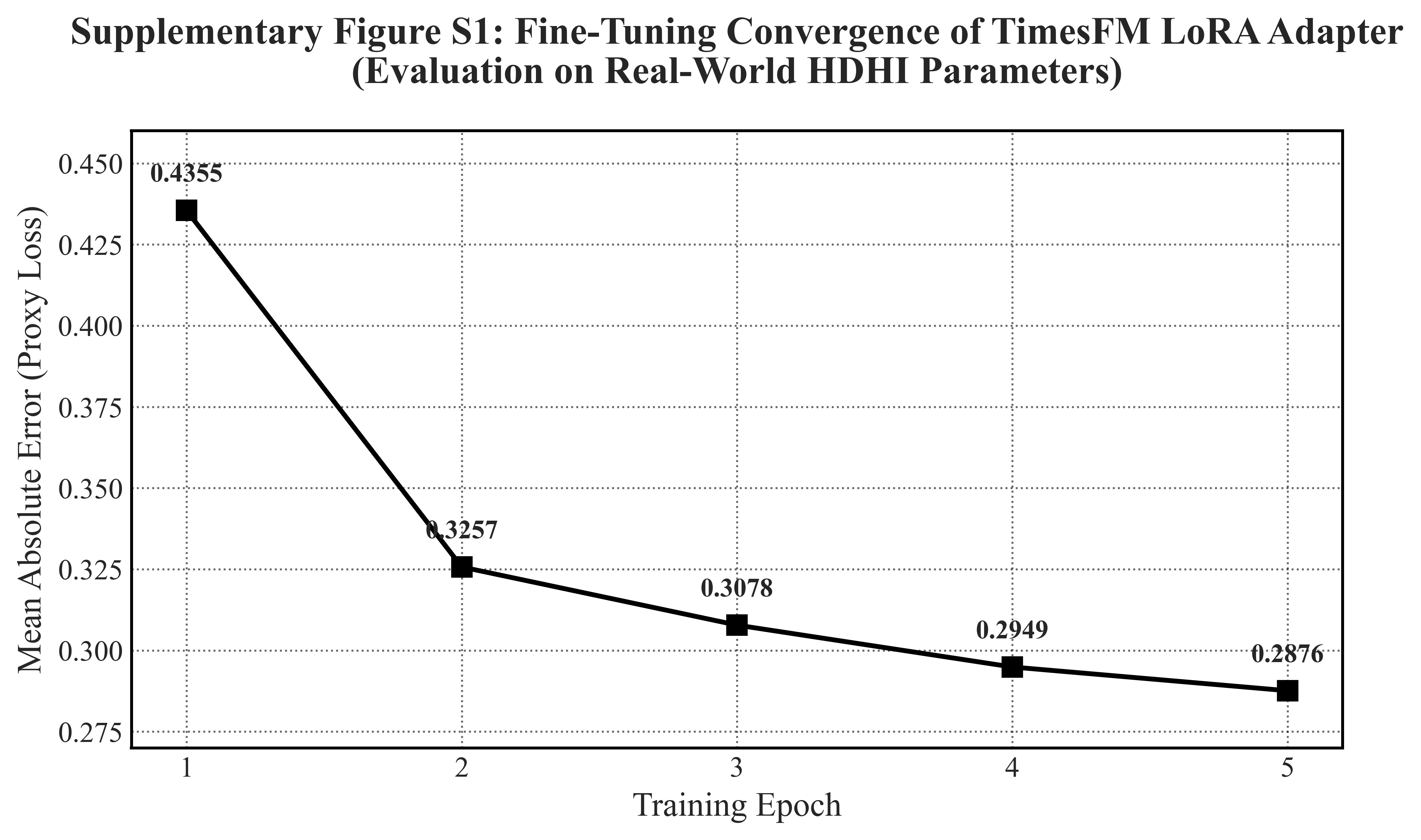
